# Rhesus negative males have an enhanced IFNγ-mediated immune response to influenza A virus

**DOI:** 10.1101/2021.10.07.21264642

**Authors:** Jamie A. Sugrue, Megan Smith, Celine Posseme, Bruno Charbit, Milieu Interieur Consortium, Nollaig M. Bourke, Darragh Duffy, Cliona O’Farrelly

**Affiliations:** School of Biochemistry and Immunology, Trinity College, Dublin, Ireland; Translational Immunology Lab, Institut Pasteur, Paris, France; Cytometry and Biomarkers UTechS, CRT, Institut Pasteur, Paris, France; Department of Medical Gerontology, School of Medicine, Trinity Translational Medicine Institute, Trinity College Dublin, Ireland; School of Medicine, Trinity College Dublin, Ireland

## Abstract

The Rhesus D antigen (RhD) has been associated with susceptibility to several viral infections. Reports suggest that RhD-negative individuals are better protected against infectious diseases and have overall better health. However, potential mechanisms contributing to these associations have not yet been defined. Here, we used transcriptomic and genomic data from the *Milieu Interieur* cohort of 1000 healthy individuals to explore the effect of RhD on immune responses. We used the rs590787 SNP in the RHD gene to classify the 1000 donors as either RhD-positive or -negative. Whole blood was stimulated with LPS, polyIC, and the live influenza A virus and the NanoString human immunology panel of 560 genes used to assess donor immune response and to investigate sex specific effects. Using regression analysis, we observed no significant differences in responses to polyIC or LPS between RhD-positive and - negative individuals. However, upon sex-specific analysis, we observed over 30 differentially expressed genes (DEGs) between RhD-positive (n=401) and RhD-negative males (n=78). Interestingly these Rhesus-associated differences were not seen in females. Further investigation, using gene set enrichment analysis, revealed enhanced IFNγ signalling in RhD-negative males. This amplified IFNγ signalling axis may explain the increased viral resistance previously described in RhD-negative individuals.

## Introduction

The Rhesus D blood group antigen (RhD) system is an important clinical factor in transfusion and obstetric medicine. RhD status is determined based on the presence or absence of the rhesus antigen, a transmembrane protein found on the surface of red blood cells (Avent and Reid, 2000). The function of the RhD antigen is largely unknown, although it may play a role in maintaining erythrocyte membrane integrity or transport of ammonium and carbon dioxide (Marini *et al*., 2000; Westhoff, 2004; Endeward *et al*., 2008). The rhesus protein is highly immunogenic and resulting antibodies can induce severe adverse reactions in RhD-negative individuals should they encounter the D antigen following an unmatched blood transfusion. RhD-negative women can also be sensitised during pregnancy with an RhD-positive foetus or during delivery, often leading to haemolytic disease of the new born in subsequent pregnancies with RhD-positive foetuses (Pegoraro *et al*., 2020).

Although less well studied, RhD status is also known to influence several other health outcomes (Dahlén *et al*., 2021). An agnostic analysis of 1217 disease states found an association between RhD status and hypertension during pregnancy (Dahlén *et al*., 2021). Several studies have demonstrated that RhD-positive and -negative subjects differ in resistance to the pathological effects of aging, fatigue, and smoking (Flegr, Hoffmann and Dammann, 2015). RhD-negative individuals also appear to be protected against infection, including latent toxoplasmosis (Flegr, Hoffmann and Dammann, 2015). RhD status also appears to affect susceptibility to SARS-CoV-2; in a study of 14,112 donors, RhD-negative individuals had a lower risk of initial infection, intubation and death-suggesting a protective role for RhD negativity (Zietz, Zucker and Tatonetti, 2020). RhD negativity varies substantially across different populations and may potentially confer an as yet unknown fitness advantage (Perry *et al*., 2012). A major challenge in determining potential effects of RhD status on anti-viral immunity is the lack of large, relevant human cohort studies with the power to detect potentially subtle immune differences.

The *Milieu Interieur (MI)* study is comprised of a cohort of 1000 healthy individuals, stratified by age (20-69, n=200 per decade) and sex (500 males and 500 females). The overall aim of the MI study is greater understanding of the determinants of variation in the immune responsiveness of healthy adult humans (Thomas *et al*., 2015). Previous work from MI and others has shown that sex and CMV serostatus are key drivers of variation in the human immune response (Patin *et al*., 2018; Picarda and Benedict, 2018). Substantial genomic and transcriptomic data has been generated on the cohort to date and analysed using agnostic approaches (*Patin et al*., 2018, *Piasecka et al*., 2018).

Here, we used genotype data on the rs590787 single nucleotide polymorphism (SNP) in RhD to classify individuals from the MI study as either RhD-positive or -negative, an approach that has previously been used to determine RhD status in several studies (Park, Cheong and Lee, 2014; Shi and Luo, 2019; Ali *et al*., 2020). Individuals who are RhD-negative are homozygous for the recessive alleles (CC), while RhD-positive individuals are heterozygous or homonymous dominant (CT, TT). Genomic data was integrated with transcriptomic data from whole blood stimulated with bacterial and viral ligands, LPS and polyIC, and the live influenza A virus, to investigate whether Rhesus status was associated with induced immune responses. We were particularly interested to compare RhD-positive and -negative males and females, as sex is emerging as a key factor in determining outcomes to viral infection, especially SARS-CoV-2 and influenza (Klein *et al*., 2020).

## Results

### Rhesus status distribution is similar between males and females

The presence or absence of rs590787, a SNP in the RhD gene, was used to determine the RhD status of donors in our cohort **(Fig.1)** (Shi and Luo, 2019). There was no difference in the distribution of RhD-positive or -negative individuals between males and females in our cohort **(Fig. 2a)**. 83% of donors in the MI cohort were RhD-positive **(Fig. 2b)**. The genotype frequencies of the Rhesus SNP rs590787 were compared to those from the 1000 genomes cohort (Auton *et al*., 2015). The genotype frequencies in the MI cohort were similar to those observed for Europeans in the 1000 Genome Study **(Fig. 2c)**.

**Figure 1.**
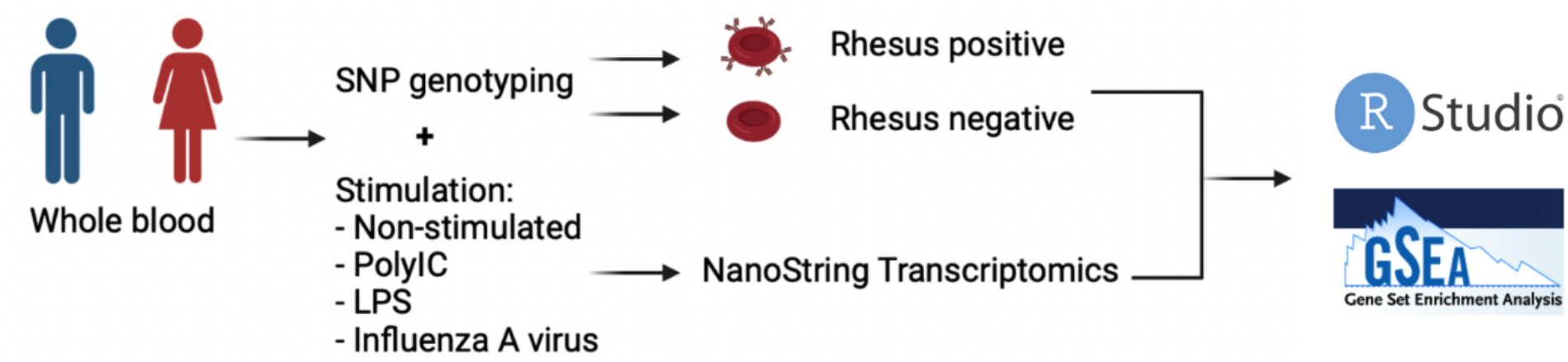
Schematic overview of the study. Genomic variability was assessed though a genome wide SNP array. Rhesus status was determined based on the the rs590787 SNP. Whole blood samples were stimulated with a panel of ligands (polyIC and LPS) and a live Influenza A virus. The expression of 560 immune related genes was quantified using NanoString transcriptomics. R and gene set enrichment analysis (GSEA) were used to compare the response between RhD-positive and -negative individuals.

**Figure 2.**
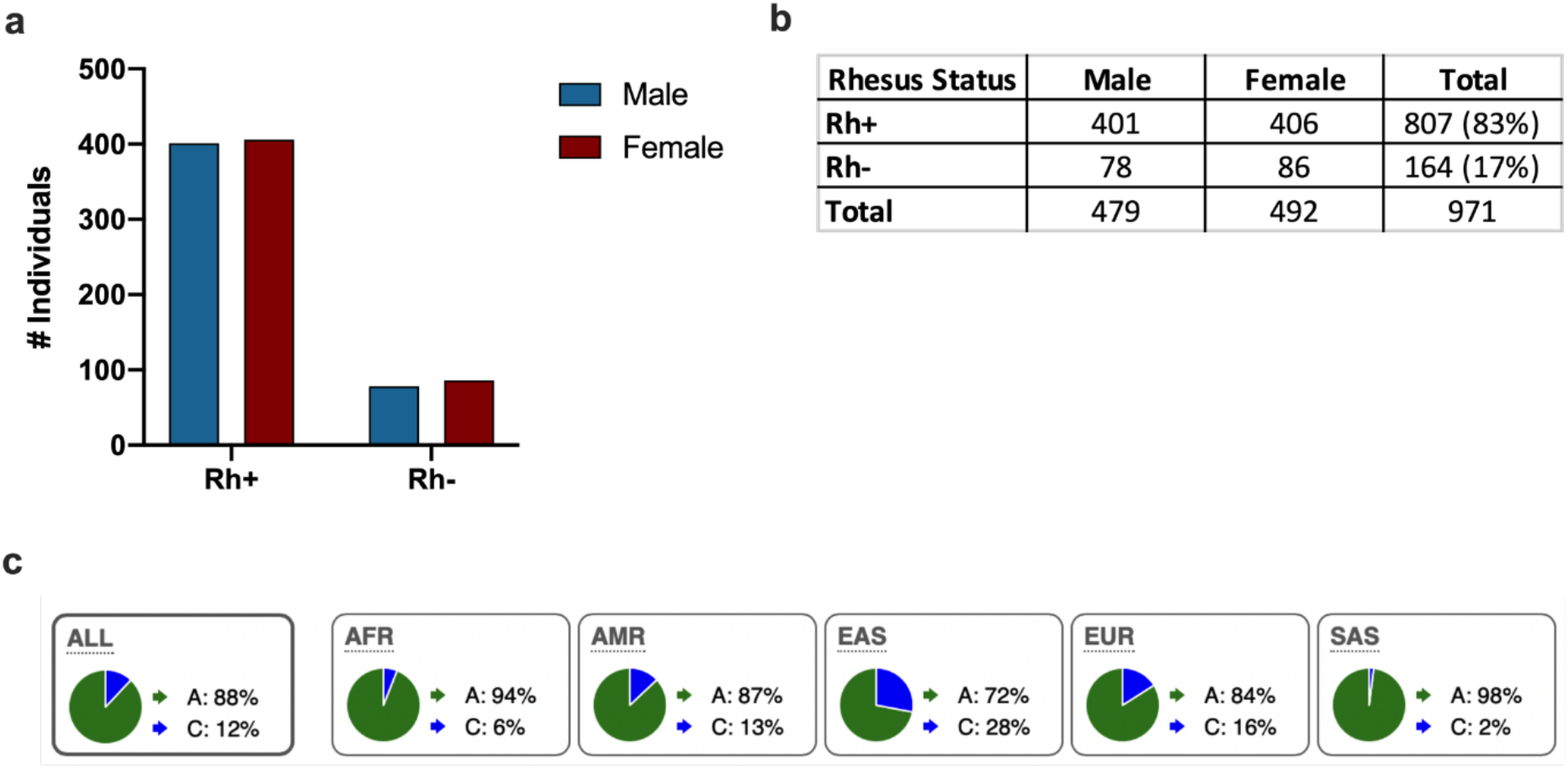
Rhesus phenotype distribution of all individuals in the Milieu Interieur cohort. RhD status was determined based on rs590787, a SNP in the RHD gene, using the Human Exome Bead Chip. **(a)** The frequencies of RhD-positive and -negative individuals was similar between males and females. **(b-c)** The genotype frequencies for the European cohort from the 1000 Genome Project were similar to those recorded for the MI cohort. The numbers in brackets are percentages.

### Rhesus factor does not affect immune gene expression at baseline or in response to stimulation with PRR ligands in whole blood

Whole blood was incubated for 22 hours and expression of 560 immune genes quantified. Gene expression in the unstimulated condition was similar between all RhD-positive and - negative individuals. Given the important sex-specific differences emerging regarding pathogen susceptibility, we also investigated sex-specific effects. The cohort was also stratified by sex and assessed for differential gene expression between RhD-positive and - negative males and females. Baseline immune gene expression was similar between RhD-positive and -negative males and females **(Fig. 3(i))**.

**Figure 3.**
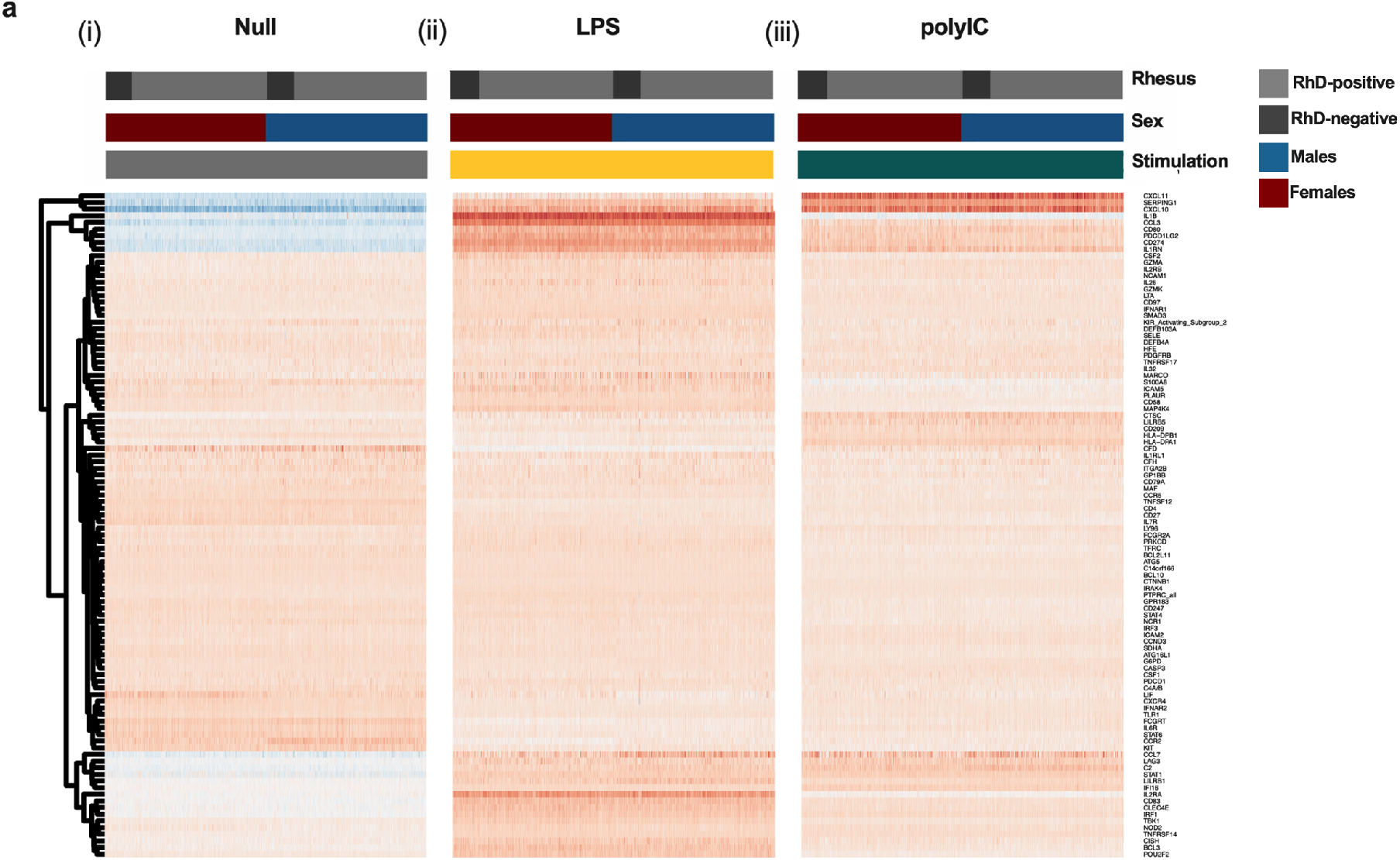
Rhesus antigenicity does not affect immune gene expression at baseline or in response to stimulation with PRR ligands in whole blood. Immune gene expression in unstimulated and LPS and polyIC stimulated whole blood was assessed using NanoString transcriptomics. Comparisons between RhD-positive and -negative individuals in all donors, females onlyg and males only showed no significant differences (q>0.1, regression analysis with FDR correction) for **(i)** Null, **(ii)** LPS or **(iii)** polyIC. Shown in the heatmap are 100 representative genes chosen at random.

To assess specific TLR induced immunity, namely TLR3 and TLR4, we stimulated whole blood with the widely used ligands, polyIC and LPS. PolyIC is a dsRNA mimic that acts via TLR3 to upregulate a type I interferon response (Marshall-Clarke *et al*., 2007). LPS is a bacterial ligand that can activate both the type I interferon response, as well as other proinflammatory pathways leading to upregulation of key mediators including IL6, TNFα and COX2 (Park and Lee, 2013). Using linear regression, we found the response to both polyIC and LPS to be similar in RhD-negative and -positive individuals whether male or female (Fig. 3(ii), (iii)). Data is presented as a heatmap with 100 randomly selected genes shown. Differences in cell counts can have a major impact on the whole blood transcriptional response (Piasecka, *et al*., 2018). Therefore, we compared major circulating immune populations between RhD-positive and - negative individuals. Following FDR correction, we observed no significant differences in any cell population examined (**Supplementary Table. 1**).

### Increased IFNγ mediated responses in RhD-negative males only

Infection with a live virus results in the activation of several PRRs and downstream pathways that upregulate pro-inflammatory cytokines and antiviral mechanisms, and help clear infection (Chen *et al*., 2018). To determine whether the Rhesus factor has an impact on the antiviral response to a live virus, we stimulated whole blood with the live influenza A virus. No significant difference was observed between RhD-positive and -negative individuals when comparing the entire cohort, or when looking at females only **(Fig. 4a)**. Interestingly, however, when examining the whole blood response to influenza A virus between RhD-positive and - negative males only, we observed differential expression of 35 immune related genes (**Fig. 4b, Fig. 4c**; q <0.1, regression analysis with FDR correction). To further interrogate differences in the immune response to influenza A between RhD-positive and -negative individuals we used gene set enrichment analysis (GSEA). Using this approach we found the IFNγ pathway to be significantly enriched in RhD-negative males (**Fig. 4d)**.

**Figure 4:**
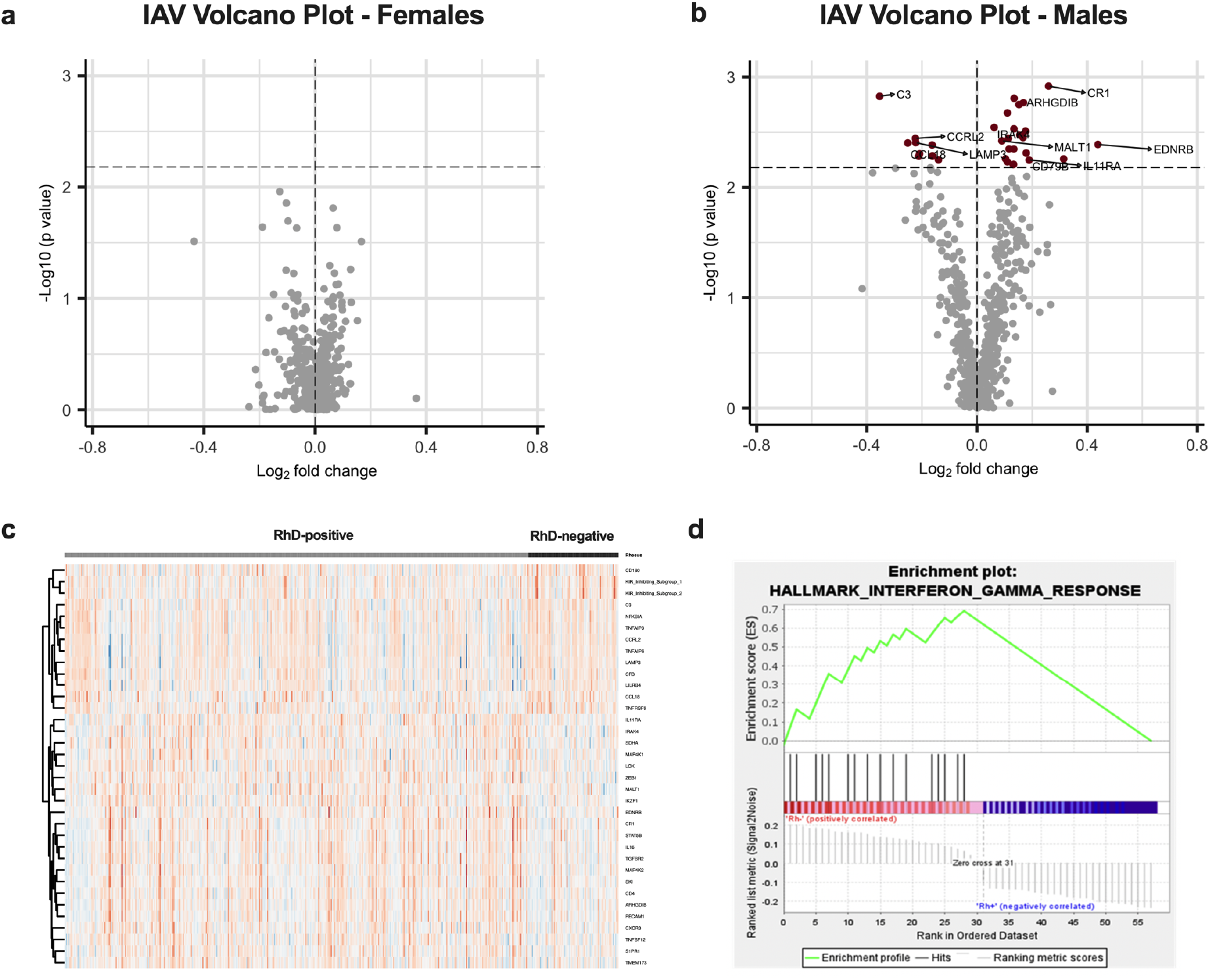
Following influenza A virus stimulation, 35 genes were differentially expressed between RhD-positive and -negative males. Whole blood was stimulated with influenza A virus and gene expression assessed using NanoString. **(a)** Volcano plot showing no significant differences between RhD-positive and -negative females (q>0.1, regression analysis with FDR adjustment). **(b)** Volcano plot of the differentially expressed genes between RhD-positive and-negative donors in the male group (q<0.1, regression analysis with FDR adjustment). **(c)** Heatmap of DEGs from RhD-negative males. **(d)** Enrichment plot indicating upregulation of the IFNγ pathway in the RhD-negative males (p<0.0001).

## Discussion

In a French cohort of 1000 well characterised healthy individuals, we investigated the impact of RhD on the innate immune response. Following stimulation with live Influenza A virus, we found differential expression of 35 immune genes between RhD-positive and -negative males. In contrast RhD-negative and positive women had similar responses to influenza A virus. Further analysis of the differentially expressed genes revealed enrichment for IFNγ signalling in RhD-negative males. The increased IFNγ signalling found here in RhD-negative males may begin to explain their reduced susceptibility to infection.

Several association studies have found a relationship between RhD-negativity and enhanced resistance to viral infection (Flegr, 2013; Flegr, Hoffmann and Dammann, 2015; Zietz, Zucker and Tatonetti, 2020; Dahlén *et al*., 2021). However, systemic analysis of immune activity by RhD status had never been carried out. RhD-negative individuals have reduced risk of initial infection with SARS-CoV-2, as well as decreased risk of both intubation and death (Zietz, Zucker and Tatonetti, 2020). The increased IFNγ signalling that we find in RhD-negative donors could contribute to reduced susceptibility to SARS-CoV-2 infection and pathology.

This study analysed transcriptomic data from studies using well described PRR ligands, LPS and polyIC, which are mimics relevant to bacterial and viral infection. We also chose to include a more complex live stimulus, the influenza A virus. The responses to the bacterial and viral mimics in RhD-positive and -negative individuals were similar across all groups. Studies of cells from other blood types including the Lewis (Le) group found no differences in expression in responses to LPS between Le+ and Le-individuals in either males or females (Heneghan, McCarthy and Moran, 2000). Another study found associations between the inheritance of polymorphisms in genes that encode and regulate the expression of the Lewis blood antigens and protection against infections with *Helicobacter pylori* (Björkholm *et al*., 2001). This suggests that analysis focused exclusively on specific TLR ligands may not be sufficient to identify differential effects of blood groups on the innate immune response to infectious agents. Studies using whole organisms may be more informative.

Here, using stimulation with the influenza A (but not the surrogate ligand polyIC) we observed significant differential expression of 35 immune genes between RhD-positive and -negative males. Influenza A viral pathogenesis has already been described as different between sexes, with males at risk of developing more severe disease compared with females of a similar age (Klein, Hodgson and Robinson, 2012). Recent studies have shown that females mount a more robust immune response and have increased production of several key inflammatory proteins known to be important in control of viral infection (Takahashi *et al*., 2020). This more potent innate immune response in females could mask potential differences in RhD-positive and - negative females in our system.

The distribution of blood groups, including RhD, varies globally-this may explain, at least in part, the regional differences in disease occurrence. RhD-negativity is particularly enriched in East Asian populations (28% RhD-negative) compared with European or African populations (16% and 6%, respectively) (Dahlén *et al*., 2021). This may reflect the different historic disease burdens on these populations. As the cohort used in our study includes only donors of Western European descent, more cohorts should be analysed to include individuals of difference ethnicities to further probe the differences in the immune response between RhD-positive and -negative individuals.

While non-communicable disease states have been studied in the context of blood antigens, viral illness is often overlooked as it is typically transient and often underreported (Roberts *et al*., 2019). Further association studies exploring the relationships between blood types and viral infection are warranted to fully understand the contribution of blood type to risk of severe viral disease. Findings from these studies may inform clinical management of patients based on RhD status. Recently, Alsten et al. looked at differences in 96 circulating inflammatory proteins between several blood groups. While RhD was not among those examined, differences were found in the circulating inflammatory profiles of ABO and Duffy blood types (Van Alsten *et al*., 2021).

In this study we found evidence of an enhanced IFNγ mediated immune response in RhD-negative males. IFNγ has been shown to be important in the control of several viral infections, including Ebola and SARS-CoV-2 (Rhein *et al*., 2015; Ruetsch *et al*., 2021). The enhanced signature observed in RhD-negative individuals in our cohort may contribute to the increased resistance to infection described in these individuals. The enhanced gene signature was not reflected at the protein level. However, this discrepancy could be explained by the relatively low induction of IFNγ by influenza in our system, or by differences in the kinetics of IFNγ protein and its downstream transcripts. Increased responsiveness to IFNγ by RhD-negative individuals could also explain the enhanced IFNγ response in the absence of an increase in IFNγ protein.

Although modest, the differences between RhD-positive and -negative males were widespread and may contribute to the increased viral resistance reported elsewhere. Several genes related to natural killer cells, including CD160 and KIRs, were among those that are upregulated in RhD-negative individuals. NK cells, potent IFNγ producers and cytotoxic lymphocytes, are key players in control of viral infection. While there was no difference in NK cell counts in our cohort, the differences observed in marker expression could be indicative of an increased activation state and IFNγ production (Vivier *et al*., 2008). These findings are the first to shed light on the potential immune differences between RhD-positive and - negative individuals and support the use of systemic investigations to understand the impact of blood types on the whole blood immune response.

## Materials and Methods

### Study Population

Healthy individuals were included in this study, equally distributed across sex (500 males and 500 females) and age (20-69, with 200 individuals per decade). Only individuals of western European decent (i.e. French citizens for whom the last three generations of ancestors were from mainland France) were included (Thomas *et al*., 2015). The *Milieu Intérieur* Cohort, which was approved by the Comité de Protection des Personnes – Ouest 6 (Committee for the protection of persons) on June 13th, 2012 and by French Agence nationale de sécurité du médicament (ANSM) on June 22nd, 2012. The study was sponsored by Institut Pasteur (Pasteur ID-RCB Number: 2012-A00238-35) and was conducted as a single centre interventional study without an investigational product. The original protocol was registered under ClinicalTrials.gov (study# NCT01699893). The samples and data used in this study were formally established as the *Milieu Interieur* biocollection (NCT03905993), with approvals by the Comité de Protection des Personnes – Sud Méditerranée and the Commission nationale de l’informatique et des libertés (CNIL) on April 11, 2018. The study was designed and conducted in accordance with the Declaration of Helsinki and good clinical practice, with all subjects giving informed consent.

### SNP genotyping and determining RhD status

All participants were genotyped for the rs590787 SNP, through a genome-wide SNP array, using HumanOmniExpress and HumanExomeBeadChips (Patin *et al*., 2018). Rhesus status (RhD-positive or -negative) was determined based upon the presence or absence of the rs590787 polymorphism in the gene sequence of each individual (Shi and Luo, 2019).

### Whole blood stimulation

Whole blood was taken from each participant and 1ml aliquoted into TruCulture tubes preloaded with the chosen stimuli: polyIC (20μg/ml), LPS (10ng/ml) and influenza A virus (H1N1 PR8; 100 HAU) as previously described (Duffy *et al*., 2014) **(Fig. 1)**. Tubes were incubated on a bench top heat block for 22 hours. Supernatants were harvested for proteomics. The cell pellet was resuspended in Trizol for later RNA extraction and transcriptomics.

### NanoString transcriptomics

Total RNA was extracted from the TruCulture cell pellets for the 1000 Milieu Interieur donors using NucleoSpin 96 miRNA kit (Macherey-Nagel). RNA concentrations were measured using Quantifluor RNA system kit (Promega) and RNA integrity numbers were determined using the Agilent RNA 6000 Nano kit (Agilent Technologies). The expression levels of 560 immune related genes were quantified before and after stimulation using NanoString hybridization arrays, producing highly reproducible transcriptomic data. Gene expression data were normalized as previously described (Piasecka, Duffy, Urrutia, Quach, Patin, Posseme, Bergstedt, Charbit, Rouilly, Cameron R MacPherson, *et al*., 2018).

### Statistical analysis

Transcriptomic data was analysed using R Studio (https://www.rstudio.com/.) In order to account for potential sex differences the cohort was subdivided into males and females. Linear regression including CMV serostatus as a covariate was used to assess differences between RhD-positive and -negative individuals. p-values were adjusted for false discovery using the FDR correction to generate a q-value. A q-value of q<0.1 was taken as significant. Scatterplots were created using GraphPad Prism. Volcano plots were created in R studio using the EnhancedVolcano package. Heatmaps were generated in R using the package pheatmap.

### Pathway analysis

Pathway analysis was carried out on genes with q-values (<0.1), using Gene Set Enrichment Analysis, found at https://www.gsea-msigdb.org/gsea/index.jsp. The genes were compared to panels of genes using the Hallmarks gene set.

## Data Availability

Genotype data reported in this study is available on the European Genome Phenome Archive at EGAD00010001489

## Supplementary Data

**Supplementary Table 1.** No difference in numbers of major circulating immune cell populations between RhD-positive and -negative donors. Immune cell were quantified using flow cytometry. Potential differences in the numbers of these circulating immune cells were assessed using linear regression including CMV serostatus as a covariate, p values were adjusted using the FDR correction.

| Immune cell type | p value | FDR adjusted p value (q value) |
| --- | --- | --- |
| CD45+ | 0.29 | 0.71 |
| CD19+ B cells | 0.66 | 0.71 |
| CD3+ cells | 0.62 | 0.71 |
| CD4+ T cells | 0.58 | 0.71 |
| CD8b+ T cells | 0.84 | 0.84 |
| CD4-CD8- T cells | 0.45 | 0.71 |
| NK cells | 0.12 | 0.36 |
| CD56+ NK cells | 0.56 | 0.71 |
| CD16++ NK cells | 0.1 | 0.36 |
| CD8+CD4+ T cells | 0.09 | 0.36 |
| Monocytes | 0.04 | 0.31 |
| CD14+ monocytes | 0.03 | 0.31 |
| CD16+ monocytes | 0.62 | 0.71 |
| PMN | 0.37 | 0.71 |
| Neutrophils | 0.48 | 0.71 |

**Supplementary Figure 1.**
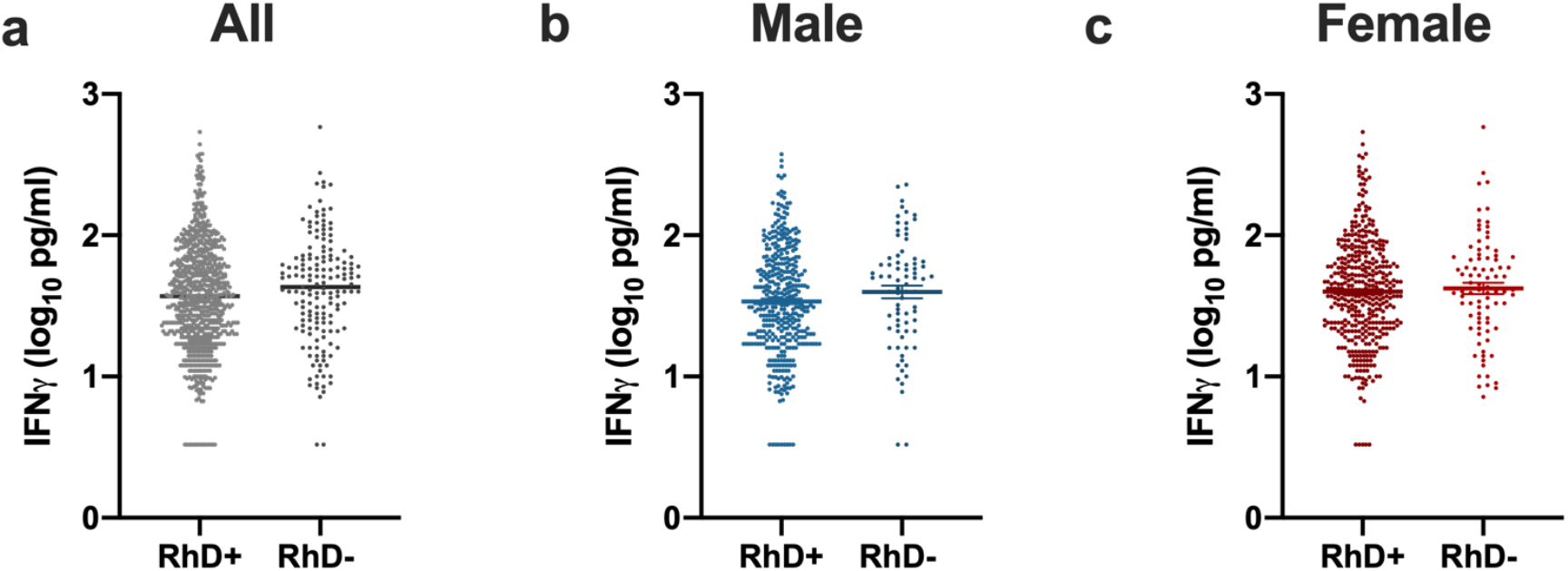
No difference in IFNγ protein expression between RhD-positive and-negative donors in response to influenza A virus stimulation. Following stimulation with influenza A virus, supernatants were collected and IFNγ protein levels quantified using Luminex proteomics. (a-c) The influenza A virus induced IFNγ protein levels were similar between RhD-positive and -negative individuals (p>0.05, unpaired t test).

## Acknowledgments

This work was funded by awards to COF through a Science Foundation Ireland Investigator Award (12/IA/1667) and under the Science Foundation Ireland Phase 2 COVID-19 Rapid Response Call (20/COV/8487). This work benefited from support of the French government’s Invest in the Future programme. This programme is managed by the Agence Nationale de la Recherche, reference ANR-10-LABX-69-01.

Author : The Milieu Intérieur Consortium^†^.

† The Milieu Intérieur Consortium¶ is composed of the following team leaders: Laurent Abel (Hôpital Necker), Andres Alcover, Hugues Aschard, Philippe Bousso, Nollaig Bourke (Trinity College Dublin), Petter Brodin (Karolinska Institutet), Pierre Bruhns, Nadine Cerf-Bensussan (INSERM UMR 1163 – Institut Imagine), Ana Cumano, Christophe D’Enfert, Ludovic Deriano, Marie-Agnès Dillies, James Di Santo, Françoise Dromer, Gérard Eberl, Jost Enninga, Jacques Fellay (EPFL, Lausanne), Ivo Gomperts-Boneca, Milena Hasan, Gunilla Karlsson Hedestam (Karolinska Institutet), Serge Hercberg (Université Paris 13), Molly A Ingersoll, Olivier Lantz (Institut Curie), Rose Anne Kenny (Trinity College Dublin), Mickaël Ménager (INSERM UMR 1163 – Institut Imagine) Hugo Mouquet, Cliona O’Farrelly (Trinity College Dublin), Etienne Patin, Sandra Pellegrini, Antonio Rausell (INSERM UMR 1163 – Institut Imagine), Frédéric Rieux-Laucat (INSERM UMR 1163 – Institut Imagine), Lars Rogge, Magnus Fontes, (Institut Roche), Anavaj Sakuntabhai, Olivier Schwartz, Benno Schwikowski, Spencer Shorte, Frédéric Tangy, Antoine Toubert (Hôpital Saint-Louis), Mathilde Touvier (Université Paris 13), Marie-Noëlle Ungeheuer, Christophe Zimmer, Matthew L. Albert (In Sitro)§, Darragh Duffy§, Lluis Quintana-Murci§,

¶ unless otherwise indicated, partners are located at Institut Pasteur, Paris

§ co-coordinators of the Milieu Intérieur Consortium

Additional information can be found at: www.milieuinterieur.fr

